# Tracheal Eccentricity is Associated with Respiratory Morbidity and Mortality in Chronic Obstructive Pulmonary Disease

**DOI:** 10.64898/2026.10.14.26361560

**Authors:** Stéphanie M. Aguilera, Venkata L. Sthanam, Muhammad F.A. Chaudhary, Arie Nakhmani, Joseph M. Reinhardt, Surya P. Bhatt, Sandeep Bodduluri

## Abstract

**Background:** Chronic obstructive pulmonary disease (COPD) is characterized by airway remodeling, which may affect tracheal morphology. However, the relationship between tracheal morphometry and COPD severity, quality of life, functional capacity, and mortality remains poorly understood.

**Objective:** To investigate the associations between CT-derived tracheal lumen eccentricity and clinical outcomes in COPD.

**Methods:** We analyzed data from 8,383 participants in the multicenter COPDGene study. Tracheal lumen eccentricity (ε) was calculated at each cross-section using the major and minor axes lengths and deriving final value using the arithmetic mean across all tracheal slices, where ε = 0 represents a circle and ε > 0 indicates an ellipse. Baseline assessments included spirometry measurements, 6-minute walk distance (6MWD), St. George’s Respiratory Questionnaire (SGRQ), and the modified Medical Research Council (mMRC) dyspnea score. Longitudinal assessments included annualized lung function change and all-cause mortality. Multivariable models were used to evaluate associations between tracheal eccentricity and clinical outcomes.

**Results:** Tracheal eccentricity increased with severity of airflow obstruction (0.60±0.07, 0.61±0.07, 0.62±0.07, 0.64±0.08, and 0.64±0.08 in GOLD stages 0 through 4, respectively). Higher tracheal eccentricity was independently associated with lower FEV_1_ (β = −0.30 L, 95% CI: −0.47 to −0.14, p<0.001), lower FEV_1_/FVC (β = −0.06, 95% CI: −0.09 to −0.04, p<0.001), lower 6MWD (β = −15.7 m, 95% CI: −25.2 to −6.2, p=0.001), higher SGRQ score (β = 16.9, 95% CI: 11.2 to 22.5, p<0.001), higher mMRC dyspnea score (β = 0.4, 95% CI: 0.1 to 0.6, p=0.007), and accelerated lung function decline (β = −23.8 ml/year, 95% CI: −2.48 to −45.13, p=0.029). Increased tracheal eccentricity is associated with increased all-cause mortality (adjusted HR: 1.81 [95% CI, 1.01-3.26]; p = 0.047).

**Conclusions:** These findings suggest that tracheal morphology may serve as an imaging biomarker for COPD outcomes, supported by significant associations with lower quality of life, lung function decline, and all-cause mortality.

## Introduction

Chronic obstructive pulmonary disease (COPD) represents a critical global health challenge and is a leading cause of mortality affecting 384 million individuals worldwide (1). COPD is recognized as a heterogeneous condition with complex pathophysiology and varied symptom burden (2,3). While spirometry serves as the diagnostic standard, it is not sensitive to early and mild disease and provides limited insight into underlying structural changes. Advances in CT imaging and quantitative analysis techniques have led to extensive phenotyping of COPD by relating structure to function, and provided novel biomarkers associated with COPD pathophysiology(4–6).

Current CT-based morphological assessments of airway disease focus predominantly on medium-sized airways and parenchymal destruction (7–10). However, the prognostic potential of central airway remodeling in COPD is increasingly being recognized. Central airway morphology may offer unique insights into both local and systemic manifestations of COPD (11–13). Recent studies have highlighted the importance of airway epithelial injury and repair mechanisms in COPD pathogenesis. As such, changes in central airway morphology may reflect ongoing inflammatory processes associated with COPD (11–13).

The tracheal lumen has been observed to adopt an eccentric or oval shape in patients with COPD, deviating from the expected circular cross-sectional geometry compared to those without COPD (14). This structural alteration is likely an extension of the airway inflammation throughout the tracheobronchial tree resulting in increased collagen deposition, fibrosis, and altered extracellular matrix composition, leading to uneven thickening of airway walls and luminal narrowing across all generations (2–3). While smaller and distal airways exhibit more severe damage due to their structural vulnerability, the inflammatory processes simultaneously affect both peripheral and central airways (7–10, 15–17).

Given the structural alterations in the central airways, we hypothesize that changes in tracheal morphology may serve as a quantifiable marker of disease burden. Therefore, we introduce tracheal lumen eccentricity, defined as the quantitative deviation of tracheal cross-section from circularity, as a novel CT-derived biomarker for COPD progression and prognosis. Using a large, well-characterized cohort of current and former smokers from the COPDGene study, we investigated the relationship between tracheal eccentricity and respiratory morbidity and all-cause mortality (14,15).

## Methods

### Study Population

We analyzed data from participants enrolled in the Genetic Epidemiology of COPD (COPDGene) study, a multicenter observational cohort study designed to investigate the genetic and epidemiologic factors associated with COPD (16). Study criteria included ages 45-80 years with at least 10 pack-years smoking history for both former and current smokers and no other significant lung disease besides COPD and asthma. A small number of lifetime non-smokers were also enrolled. Pre- and post-bronchodilator spirometry were performed according to the American Thoracic Society (ATS) guidelines (17). Disease severity was classified using the Global Initiative for Chronic Obstructive Lung Disease (GOLD) staging system, ranging from Stage 0 (smokers without airflow obstruction) to Stage 4 (severe functional impairment). Metrics included the forced expiratory volume in one second (FEV_1_) and the FEV_1_/forced vital capacity (FVC) ratio. The St. George’s Respiratory Questionnaire (SGRQ) was administered to assess respiratory health-related quality of life, with higher scores indicating worse quality of life (18,19). The 6-minute walk test was conducted following standardized protocols to measure functional capacity (10). The modified Medical Research Council (mMRC) dyspnea scale was administered to assess level of activity related breathlessness, with higher scores indicating increased symptom burden (20). Baseline assessments were repeated at approximately 5 years (visit 2). Lung function change was calculated as the annualized change in FEV_1_ between visits 1 and 2. Participants with preserved ratio impaired spirometry (PRISm), classified as having reduced lung function but not meeting the spirometry definition of COPD, were excluded from this study (21). Exclusions also included unavailable airway segmentations from inspiratory CT scans. Participant selection and exclusion in this COPDGene secondary analysis are depicted in the CONSORT diagram in **Figure 1**.

**Figure 1.**
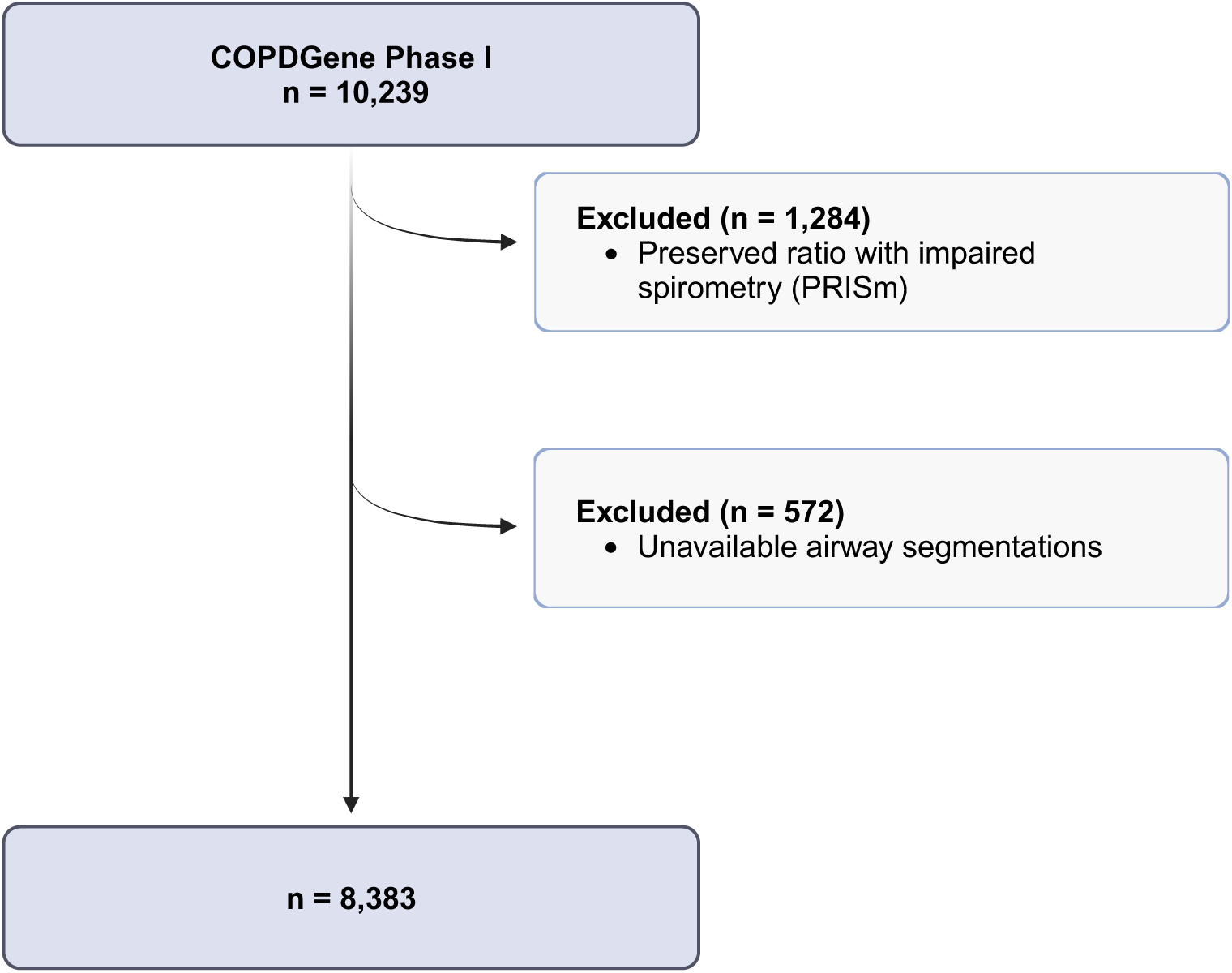
COPDGene Study CONSORT diagram.

The study was approved by the institutional review boards of all participating centers, and all participants provided written informed consent.

### CT Imaging

Participants underwent volumetric chest CT scans at full inspiration using multidetector CT scanners. Scanning protocols were standardized across all centers (16). CT images were acquired with the participant in the supine position during a single breath hold. Standard-dose scans were acquired at visit 1, adhering to scanner specific protocols and acquisition parameters previously reported (22). The images were reconstructed with the B31f kernel, with a slice thickness of 0.7 mm and a reconstruction interval of 0.5 mm. LungQ (Thirona, Netherlands) imaging suite was used to extract lungs and segment airway (**Figure 2-A**) (7,23).

**Figure 2.**
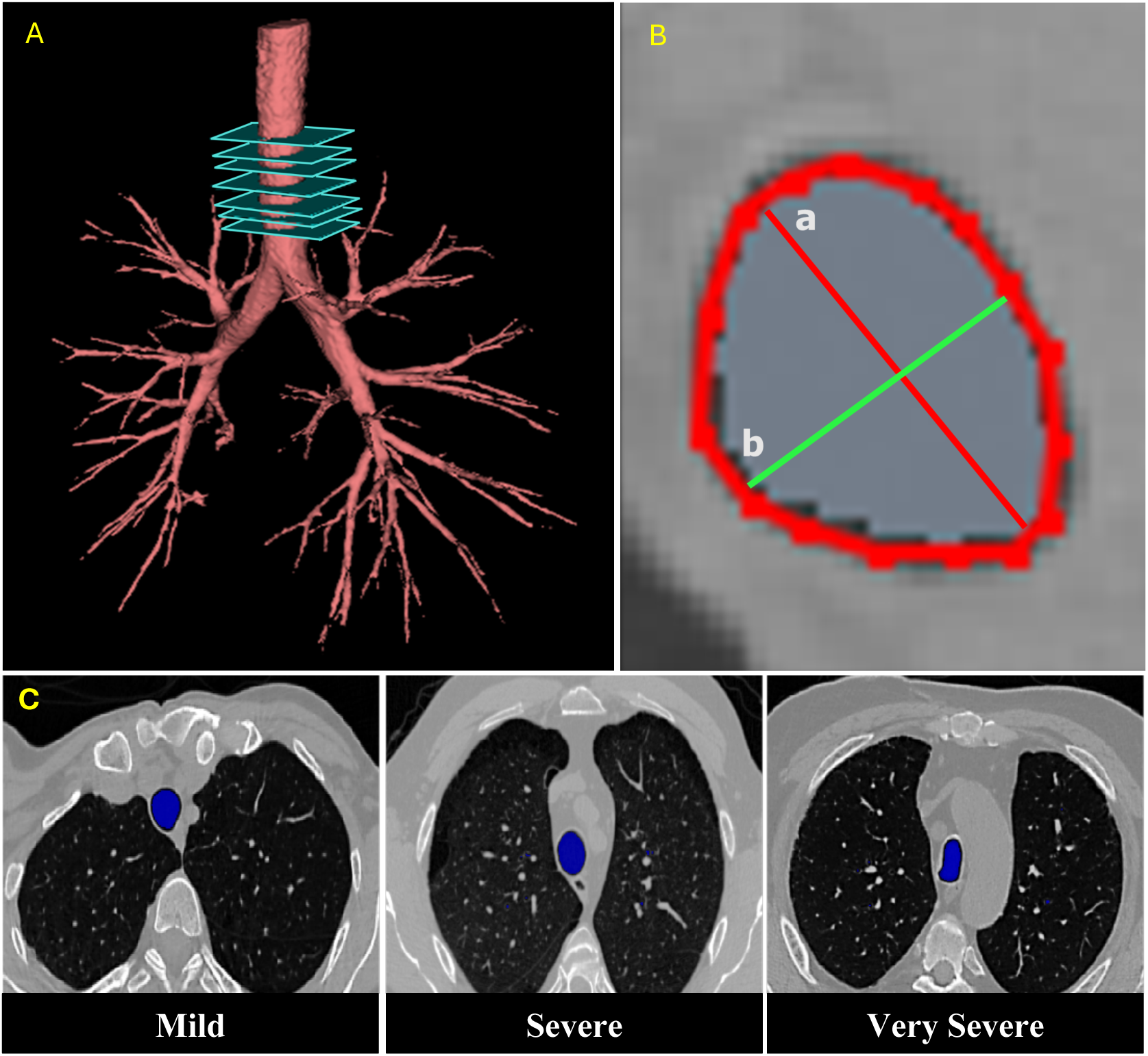
***Top Row:*** (A) Representative image of a segmented airway from a participant with GOLD stage 0. (B) Axial inspiratory CT scan cross-section with delineated trachea. Major axis (a) and minor axis (b) denote the largest and smallest diameters across the cross-section, respectively. A value of 0 corresponds to a perfect circle, while a value greater than 0 corresponds to an ellipse. Mean eccentricities greater than 0 along the trachea represent increased asymmetry between the major and minor axis lengths. ***Bottom Row:*** (C) Representative axial CT inspiratory scans of trachea among three distinct eccentricity severity quartiles in GOLD 0 subjects with eccentricity values of 0.49, 0.62, and 0.80, respectively.

### CT Metrics

Pi10, which refers to the airway wall thickness, was computed using the square root of wall area of a 10-mm lumen perimeter (24). Emphysema was quantified using the percentage lung volume with attenuation values below −950 HU (%LAA −950)(25).

### Tracheal Eccentricity

Using the segmented airway trees, cross-sectional measurements were extracted from sequential 2D slices perpendicular to the airway centerline, with reference landmarks spanning from the most superior part of the trachea to the carina. Slice-specific tracheal eccentricity (ε) was derived as 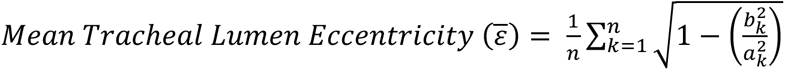 where: ***a_k_*** : major axis length at slice k, ***b_k_*** : minor axis length at slice k, ***n*** : total number of slices, using MATLAB R2023a (MathWorks, Inc.) as shown in **Figure 2-B**. Tracheal lumen eccentricity was quantified using the arithmetic mean of eccentricity across all slices. This operation yielded scalar values of range [0, 1], where ε = 0 corresponds to a perfect circle (b_k_ = a_k_), and ε greater than 0 denotes an ellipse. The eccentricity metric represents the deviation of tracheal cross-section from circularity, with higher eccentricity values closer to 1 reflecting more elliptical and asymmetric tracheal morphology.

### Statistical Analysis

Shapiro-Wilk test and QQ plots were conducted to evaluate the normality of data. The Kruskal-Wallis rank-sum test was performed to evaluate differences in mean tracheal lumen eccentricity across GOLD stages. The Jonckheere-Terpstra test was also performed to evaluate whether mean tracheal eccentricity followed a monotonic trend across GOLD stages. Associations between eccentricity and lung function metrics (FEV_1_, FEV_1_/FVC), clinical outcome measures (6-minute walk distance, SGRQ score, mMRC dyspnea score), and annualized change were tested using generalized linear regression models. Lung function models were adjusted for age, race, sex, body mass index (BMI), smoking status, pack-years of smoking, scanner type, Pi10, and %CT emphysema. Clinical outcomes measures and annualized FEV1 change models, were adjusted for all metrics previously mentioned and baseline pre-bronchodilator FEV_1_. Associations between eccentricity and all-cause mortality were evaluated using a Cox proportional hazards model, adjusting for covariates as denoted in generalized models. To quantify mortality risk associations, tracheal eccentricity was categorized into quartiles of severity, and evaluated using multivariable cox proportional hazards analyses. All covariates were evaluated for collinearity with tracheal eccentricity using variance inflation factor (VIF). Statistical analyses were performed using the R statistical software (version 4.3.1). Statistical significance was defined by two-sided p-value < 0.05.

## Results

### Participant Characteristics

8,383 smokers and non-smokers were included in the current analysis (CONSORT diagram in **Figure 1**). Participants had a mean age of 59.9 (SD 9.1) years. The cohort was comprised of 3,834 (45.7%) females and 2,600 participants (31.0%) identified as African American. Additionally, 4,230 (50.5%) were current smokers and mean (± SD) BMI was 28.4 (SD 5.9) kg/m². CT-based mean (± SD) percent emphysema was 6.93% (SD 10.21), the mean (± SD) Pi10 was 2.33 mm (SD 0.62), and mean (± SD) tracheal eccentricity was 0.61 (SD 0.08) (**Table 1).**

**Table 1:** Baseline Demographics of Participants (n = 8,383)

| **Parameters** | **Results** |
| --- | --- |
| **Age, years** | 59.9 (9.09) |
| **Sex, Female, n (%)** | 3,834 (45.7%) |
| **African American race, n (%)** | 2600 (31.0%) |
| **Body Mass Index, kg/m<sup>2</sup>** | 28.36 (5.91) |
| **Current Smokers, n (%)** | 4,230 (50.5%) |
| **Pack years of smoking** | 43.9 (25.3) |
| **FEV<sub>1</sub>, L** | 2.29 (0.95) |
| **FEV<sub>1</sub> %predicted** | 77.8 (26.8) |
| **FEV<sub>1</sub>/FVC** | 0.66 (0.17) |
| **GOLD Stage, n (%)** |  |
| **Nonsmokers:** | 106 (1.3%) |
| **0:** | 4,102 (48.9%) |
| **1:** | 748 (8.9%) |
| **2:** | 1,810 (21.6%) |
| **3:** | 1,076 (12.8%) |
| **4:** | 541 (6.5%) |
| **% Emphysema** | 6.93 (10.21) |
| **Pi10, mm** | 2.33 (0.62) |
| **Tracheal Eccentricity** | 0.61 (0.08) |

All values expressed as mean (SD) unless specified otherwise.
FEV<sub>1</sub>: Forced Expiratory Volume in the first second.
FEV<sub>1</sub> % predicted estimated according to National Health and Nutrition Examination Survey III reference equations.
FVC: Forced Vital Capacity
CT: Computed tomography

### Mean Tracheal Lumen Eccentricity across GOLD Stages

There was a statistically significant difference in tracheal eccentricity across GOLD stages (Kruskal-Wallis Rank test, p < 0.001, Jonckheere-Terpstra test, p < 0.001) as illustrated in **Figure 3**. There was a progressive increase in mean tracheal eccentricity from GOLD 0 (0.60 ± 0.07) to GOLD 4 (0.64 ± 0.08).

**Figure 3.**
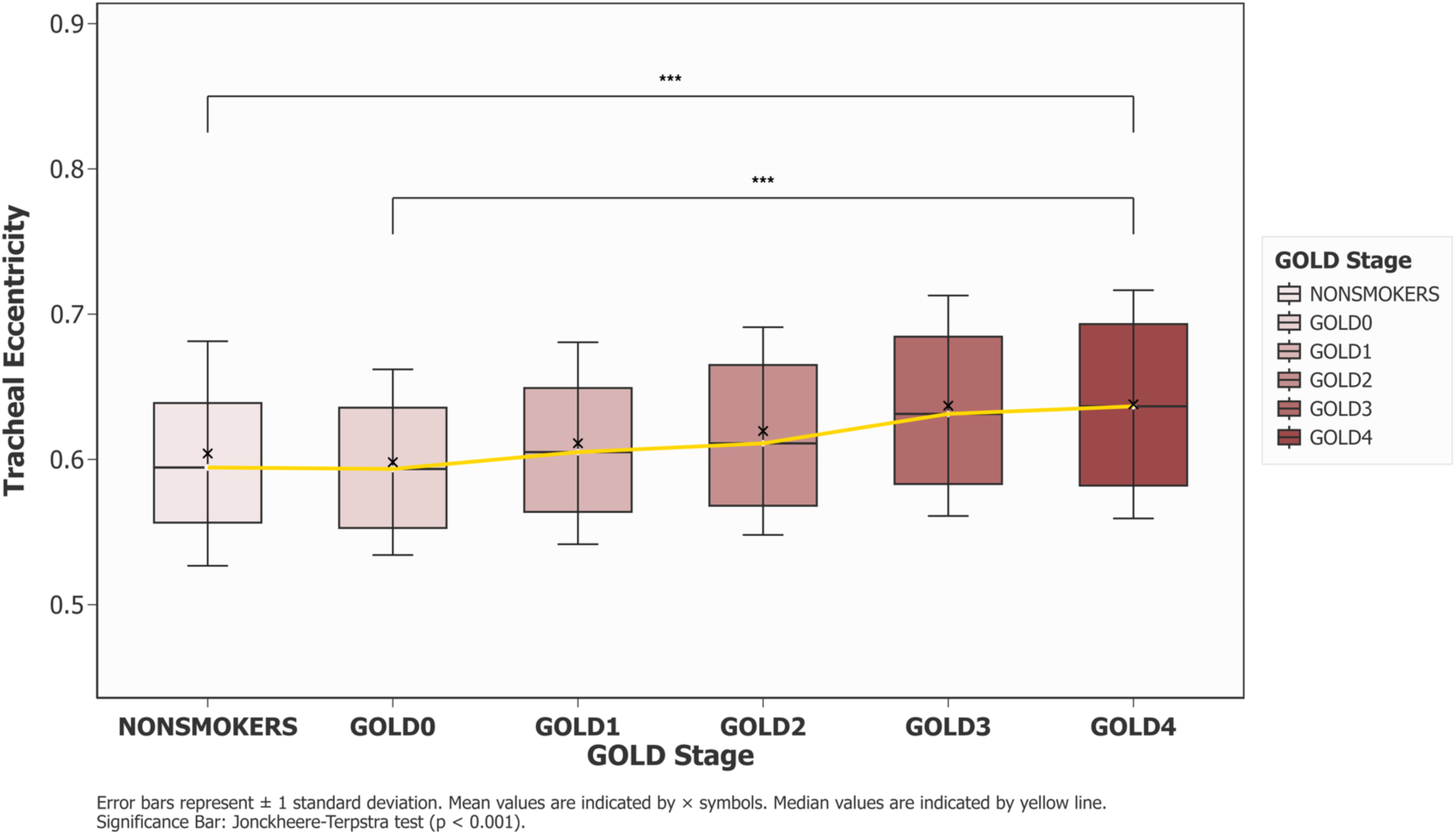
Box-plots demonstrating monotonic increasing trend of mean tracheal eccentricity observed across GOLD stages (Jonckheere-Terpstra test, p < 0.001). GOLD: Global Initiative for Obstructive Lung Disease.

### Associations Between Tracheal Eccentricity and Clinical Outcomes

Tracheal lumen eccentricity was significantly associated with clinical outcomes (**Table 2**). No concerning collinearity was detected with all VIF values below 4 (highest VIF = 3.56 for FEV_1_). Additionally, correlation coefficients between trachea eccentricity and covariates were low (all r < 0.21). Greater eccentricity correlated with lower lung function, as demonstrated by lower FEV_1_ (β = −0.30 L; 95% CI: −0.47 to −0.14; p < 0.001) and FEV_1_/FVC (β = −0.06; 95% CI: −0.09 to −0.04; p < 0.001). Greater eccentricity was associated with reduced 6-minute walk distance (β = −15.7 meters; 95% CI: −25.2 to −6.2; p = 0.001), higher SGRQ score (β = 16.9; 95% CI: 11.2 to 22.5; p < 0.001) and higher mMRC dyspnea score (β = 0.4, 95% CI: 0.1 to 0.6, p=0.007), after adjusting for age, race, sex, BMI, smoking status, pack-years of smoking, scanner type, Pi10, %CT emphysema and baseline FEV_1_. Higher eccentricity was associated with accelerated FEV_1_ decline (β = −23.8 mL/year; 95% CI: −2.5 to −45.1; p = 0.029) and with greater all-cause mortality (HR = 1.81, 95% CI: 1.01 to 3.26, p = 0.047) after adjusting for age, race, sex, BMI, smoking status, pack-years of smoking, scanner type, Pi10, %CT emphysema, and baseline FEV_1_.

**Table 2.** Multivariable Association Analysis of Tracheal Eccentricity and Clinical Outcomes.

| | $\beta$ | 95% CI | p-value |
| --- | --- | --- | --- |
| **FEV<sub>1</sub> (L)**<sup>1</sup> | -0.30 | -0.47 to -0.14 | < 0.001 |
| **FEV<sub>1</sub>/FVC**<sup>1</sup> | -0.06 | -0.09 to -0.04 | < 0.001 |
| **6 Minute Walk Distance**<sup>1,2</sup><br>**(meters)** | -15.7 | -25.2 to -6.2 | 0.001 |
| **St. George's Respiratory**<sup>1,2</sup><br>**Questionnaire (SGRQ)** | 16.9 | 11.2 to 22.5 | < 0.001 |
| **mMRC Dyspnea Score**<sup>1,2</sup> | 0.4 | 0.1 to 0.6 | 0.007 |
| | $\beta$ | 95% CI | p-value |
| **Change in FEV<sub>1</sub>**<sup>1,2</sup><br>**(mL/year)** | -23.8 | -2.5 to -45.1 | 0.029 |
|  | HR | 95% CI | p-value |
| **All-Cause Mortality**<sup>1,2</sup> | 1.81 | 1.01 to 3.26 | 0.047 |

<sup>1</sup> Adjusted for age, race, sex, BMI, smoking status, pack-years of smoking, scanner type, Pi10, and %CT emphysema.
<sup>2</sup> Additional adjustments for baseline postbronchodilator FEV<sub>1</sub>.

#### Tracheal Eccentricity Quartiles and Mortality

We divided tracheal eccentricity measurements into quartiles. **Figure 2-C** illustrates the deformation of tracheal morphology across different eccentricity quartiles. The mild quartile (0.356 to 0.561, mean ± SD: 0.52 ± 0.03) represents minimal tracheal deformation, with greater tracheal deformation with moderate (0.561 to 0.604, mean ± SD: 0.58 ± 0.01), severe (0.604 to 0.654, mean ± SD: 0.63 ± 0.01), and severe tracheal eccentricity (0.654 to 0.994, mean ± SD:0.71 ± 0.05) quartiles.

Reduced FEV_1_ was observed across tracheal eccentricity quartiles (mean ± SD: 2.36 L ± 0.89, 2.34 L ± 0.92, 2.28 L ± 0.96, and 2.12 L ± 1.03 for the mild, moderate, severe, and very severe quartiles, respectively). Similar decreasing trend was observed in FEV_1_/FVC across quartiles (mean ± SD: 0.69 ± 0.15, 0.68 ± 0.15, 0.65 ± 0.17, and 0.59 ± 0.18 for the mild, moderate, severe, and very severe quartiles, respectively). CT-based percent emphysema increased across tracheal eccentricity quartiles (mean ± SD: 5.13 ± 9.09, 5.46 ± 8.68, 7.24 ± 10.60, and 9.70 ± 11.70 for the mild, moderate, severe, and very severe quartiles, respectively). A similar trend was observed for Pi10 (2.24 ± 0.58, 2.29 ± 0.62, 2.33 ± 0.60, and 2.47 ± 0.65 for mild, moderate, severe, and very severe quartiles, respectively).

Compared with participants in the lowest quartile, those with severe (adjusted HR: 1.50 [95% CI, 1.27-1.77]; p < 0.001) and very severe eccentricity (adjusted HR: 2.57 [95% CI, 2.20-3.01]; p < 0.001) demonstrated significantly higher mortality risk (**Figure 4)**. Of the 2377 reported deaths within 13 years of follow-up, 31.6% (n = 752) were reported in individuals with very severe tracheal eccentricity. Furthermore, we found that significant tracheal deformation was present even in participants with GOLD 0, with 18.6% (n = 120) showing very severe tracheal eccentricity.

**Figure 4.**
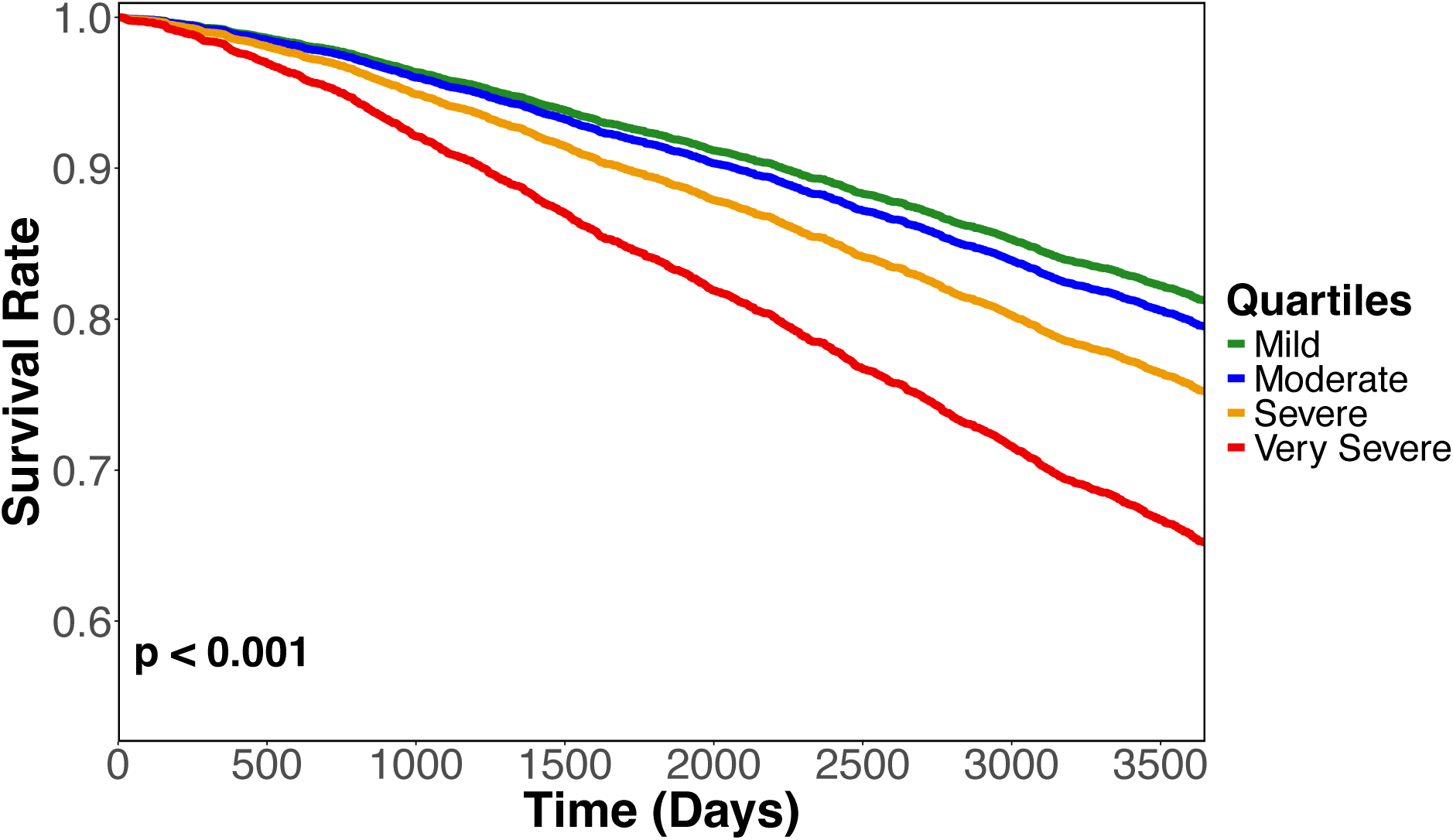
Adjusted Cox proportional hazard survival curves stratified by eccentricity quartiles with a total of 2377 deaths among patient cohort.

## Discussion

We demonstrate the significance of tracheal morphology, highlighted by alterations in tracheal eccentricity, and its associations with COPD severity and outcomes. We also observed a significant number of participants without airflow obstruction with severe tracheal eccentricity, suggesting that tracheal morphological changes may, in some cases, precede the development of airflow limitation.

Tracheal morphometry has been of considerable interest in respiratory conditions, including COPD (26–31). Classical observations such as "saber-sheath trachea” and lunate shapes have long been recognized as potential markers of disease (32–34). However, these alterations have traditionally been attributed as consequences of parenchymal damage rather than recognized as separate structural changes occurring in the tracheal wall (14,35,36). Although our findings potentially suggest tracheal abnormalities caused by epithelial-driven inflammatory and remodeling processes, the mechanisms underlying the associations observed between tracheal eccentricity and COPD outcomes remain unknown. It is plausible that some of these changes are also attributable to the influence of airway shape on airflow dynamics. In the upper airways, including the trachea, airflow is characteristically turbulent rather than laminar due to the high velocity of airflow and the irregularities in the airway lumen. Efficient gas exchange depends on the upstream transition from turbulent to laminar flow. Thus, any increase in turbulence due to tracheal irregularities could affect the mechanics of breathing (37–40). While increased tracheal eccentricity is unlikely to impact expiratory airflow resistance, the altered geometry could induce additional turbulence, thereby increasing respiratory workload. This effect may be particularly relevant for patients with already diminished respiratory reserve, where an increase in the work of breathing could contribute to adverse clinical outcomes.

Earlier CT-based studies based on the delineation of four equiangular rays (positioned 90° apart) extended from the tracheal centroid at a single mid-tracheal cross-section, yielded limited clinical significance in correlating tracheal structure with all-cause mortality (14). In contrast, our approach assesses global cross-sectional eccentricity across the trachea and offers a clinically relevant metric for evaluating tracheal morphology. Our findings are also consistent with recent studies emphasizing the importance of central airway structural abnormalities in COPD. For instance, Zhang and colleagues demonstrated the significance of upper airway structural abnormalities in facilitating early diagnosis and understanding the development of COPD (6). While their focus was on the upper airways, including the trachea, our study focused solely on the trachea, allowing for more discrete characterization. Our observations are also consistent with the work from Puerta and colleagues, who analyzed morphometric lumen irregularities in pre-COPD and mild COPD lungs and identified increased eccentricity among COPD patients in comparison to controls at end-inspiration, particularly in the trachea and first-generation bronchi (41). Additionally, we observed that there are participants with GOLD stage 0 in the severe and very severe tracheal eccentricity quartiles, showing that tracheal deformation might occur before airflow obstruction is detected through spirometry.

While our study shows significant findings about tracheal morphology and associations with clinical outcomes, our approach has inherent limitations. While our CT-based assessments provide spatial resolution in the morphometric changes of disease, they do not fully capture the dynamic nature of airway function. Furthermore, the observed associations between tracheal morphology and COPD warrant further validation particularly in how specific inflammatory remodeling processes, such as neutrophil infiltration or even mitochondrial dysfunction, are contributing to these structural alterations (36,40,42–44). Additional studies combining CT morphometry with functional imaging techniques may prove valuable in examining associations between tracheal morphology and airflow characteristics, providing additional context to the broader understanding within the development and progression of COPD.

## Conclusion

In conclusion, we showed that tracheal eccentricity is an independent predictor of COPD patient outcomes and mortality. While tracheal eccentricity increases with COPD severity, a subset of participants without airflow obstruction has severe tracheal eccentricity, suggesting that central airway remodeling may precede clinically detectable airflow limitation.

## Data Availability

All data produced in the present study are available upon reasonable request to the authors. All data produced in the present work are contained in the manuscript.

## Acknowledgements

This work was supported by NHLBI K01HL163249 (SB), NHLBI R01HL151421 (SPB & AN), U01 HL089897 and U01 HL089856. The COPDGene study (NCT00608764) is supported by grants from the NHLBI (U01HL089897 and U01HL089856), by NIH contract 75N92023D00011, and by the COPD Foundation through contributions made to an Industry Advisory Committee that has included AstraZeneca, Bayer Pharmaceuticals, Boehringer-Ingelheim, Genentech, GlaxoSmithKline, Novartis, Pfizer and Sunovion.

## Author Contributions

*Study concept and design*: Aguilera, Bhatt, and Bodduluri

*Acquisition, analysis, or interpretation of data*: All authors

*Drafting of the manuscript*: Aguilera, Bhatt, and Bodduluri

*Critical revision of the manuscript for important intellectual content*: All authors

*Statistical analysis*: Aguilera and Bodduluri

*Study supervision*: All authors

